# Change in burden of disease in UK children and young people (0-24 years) over the past 20 years and estimation of potential burden in 2040: analysis using Global Burden of Disease (GBD) data

**DOI:** 10.1101/2021.02.20.21252130

**Authors:** Joseph L. Ward, Dougal Hargreaves, Steve Turner, Russell M. Viner, On behalf of the Royal College of Paediatrics and Child Health Paediatrics 2040 Data Working Group

**Author notes:** Correspondence: Prof. Russell Viner, UCL Great Ormond St. Institute of Child Health, 30 Guilford St. London WC1N 1EH.

## Abstract

**Background:** The epidemiological transition and medical innovations have led to changes in causes of ill-health and disability by children and young people (CYP) in many wealthy countries over the past two decades. However this has not been systematically examined at a national level in the UK. Here we examined changes in disability-adjusted life-years (DALYs) by cause for 0-24 year olds by age-group.

**Methods:** We used data on DALYS by cause, sex and age-group for the UK from 1998 to 2017 from the 2017 Global Burden of Disease (GBD) study. We modified the GBD cause-hierarchy to be more relevant to paediatrics. We assessed current causes of burden in 2017 and change at cause-level for 1998-2007 and 2008-2017 by age. We then used Holt-Winters doubly exponentiated time-series models to forecast change in DALYs by age to 2040.

**Results:** In 2017, neonatal and congenital disorders were the main causes of DALYS across 0-24 year olds, with other the other large causes being anxiety and depression, endocrine and immune disorders, and lower respiratory tract infections. Total DALYS were highest amongst neonates and lowest amongst 1-9 year olds, rising with age amongst 10-24 year olds. Between 1998-2017, total DALYs fell in each age-group, with the largest falls in infants. The greatest changes in DALYS from 2008 to 2017 were falls in neonatal and congenital causes amongst infants, falls in infectious diseases and injuries in older age-groups, and rises in neonatal causes, mental health, acne and somatic symptoms in all age-groups other than infants. These patterns were forecast to continue to 2040.

**Conclusions:** We forecast falls in causes that have historically dominated disease in CYP, particularly congenital disorders, infectious diseases, cancers and injuries, representing falls in the prevalence of many infectious diseases and improvements in road safety and also improvements in survival from cancer and many congenital conditions. Forecast increases in DALYS from mental health problems, other adolescent health issues and the consequences of neonatal survival, such as neuro-disability and epilepsy, have potential implications for the training of paediatricians and workforce needs over the next two decades. The impact of the COVID-19 pandemic, climate change and changes in child poverty require further research.

## Background

The past two decades have witnessed major changes in the lives of UK children and young people (CYP) and major increases in use of health services, particularly emergency services^1^. There have also been a number of broader changes in patterns of disease in these age-groups. The reduction in burden from infectious diseases seen since the 1950s^2^ has been accelerated by changes in treatment and the introduction of new vaccines. There have been major improvements in survival of extremely premature babies, however this has resulted in increases in neurodisability amongst survivors^3^. There have also been shifts in the numbers of CYP diagnosed with autism and learning difficulties, and the prevalence of mental health problems in the general population has risen considerably^4^. These factors and others have contributed to an increase in complexity for paediatric services^5^, allied with rising parental expectations of modern medicine. Sociodemographic change may also contribute including through increasing deprivation in the CYP population^6^. There have been similar global changes in burden of disease, tracked for the last two decades by the Global Burden of Disease (GBD) study, run from the Institute of Health Metrics and Evaluation (IHME) in Seattle. GBD produces high quality estimates of ill health and injury for all most all countries of the world including detailed estimates available for each UK country^7^. GBD estimates are used routinely by public health organisations in most UK countries.

The speed of change in burden of disease amongst CYP may present challenges for health service provision, as training of professionals and models of care based upon historic patterns may not fully meet the needs of CYP today. The Royal College of Paediatrics and Child Health (RCPCH) began its Paediatrics 2040 project in 2019 (https://www.rcpch.ac.uk/work-we-do/paediatrics-2040/about), aiming to explore how changes in burden of disease and medical innovation might impact upon future models of care, workforce needs and training and support needs of paediatricians.

In this paper, we aim to understand change in burden of disease amongst CYP over the past two decades in the UK and use these estimates to forecast potential change over the twenty years to 2040, to inform Paediatrics 2040 work on models of care, workforce and training. There are a number of measures of burden of disease available to use. Healthcare activity data such as primary-care attendance or hospital admission rates are one such measure, but are biased by help-seeking factors and the organisation of the health system (e.g. access to care) and over-represent more potentially-lethal conditions and under-count less serious conditions that may cause significant burden to CYP (e.g. acne and some mental health problems). Measures that provide a more accurate picture of burden of disease at a population level include mortality rates and the impact of a condition on the loss of life-expectancy in a population, i.e. years of life lost due to premature mortality (YLLs). However, outside neonatal life, mortality amongst CYP is rare and contributes little to overall burden of disease. Measures of morbidity that are comparable across different conditions include years lived with disability (YLDs), calculated from the prevalence of a condition multiplied by pre-defined disability weights corrected for comorbidity^8^; and disability-adjusted life-years (DALYs), which are the sum of YLLs and YLDs for a cause for a location by age-group, sex, and year. DALYS provide an overall estimate of the burden related to a cause, integrating morbidity and mortality and thus reflect the contribution of each to overall burden of disease^9^. Here we use GBD DALY data for the UK to examine change in burden of disease over the last 2 decades and forecast potential trends over the next 20 years to 2040.

## Methods

Data: We used data from the GBD 2017 study, publicly available from IHME (http://www.healthdata.org/; downloaded on 10 Feb 2019). The GBD calculates YLLs and YLDs and therefore DALYS from a range of data on prevalence of and mortality from each condition by age, sex and country from a range of sources including systematic reviews of the literature, unpublished documents, survey microdata, administrative records of health encounters, registries, and disease surveillance systems. Sources are available from the GBD Data Input Sources Tool (http://ghdx.healthdata.org/gbd-2017/data-input-sources). Ages of interest for these analyses are 0-24 years as per the NHS Long Term Plan^10^ using the categories Neonatal, Postneonatal, Infants and 1-4, 5-9, 10-14, 15-19 and 20-24 year age groups. Note that infants were combined across 0-365 days for some analyses and split into neonatal and post-neonatal for others.

The GBD uses a modification of the International Classification of Disease (ICD) system that has a hierarchical structure based upon on the type of cause rather dominated by organ system as does the ICD. The GBD cause hierarchy has 4 levels of detail, with Level 3 (169 cause categories) chosen as the most suitable level for these analyses. As many Level 3 causes are little relevant for CYP (e.g. diseases of ageing, rare adult cancers) or extremely rare in the UK (many tropical diseases), we grouped causes into a modified cause-hierarchy more relevant to paediatrics and grouped some causes with very low levels of DALYS (see Appendix Table A1).

## Analyses

All analyses were undertaken using Stata 15.0 (StataCorp; College Station, TX) and visualisations were undertaken using Tableau 2020. Analyses were undertaken in 3 sequential steps, 1) outlining current burden by age and sex in the UK; 2) identifying change at cause level in the UK over the last two decades (1998-2007; 2008-2017) by age; and 3) forecasting future UK burden by cause and age to 2030 and 2040. There was extremely high autocorrelation in the time series for the top 30 causes of DALYS by age from 2008-2017 for each age (Appendix Table 2) which supports the use of exponential smoothing for forecasting. Forecasting used Holt-Winters doubly exponentiated time-series smoothing models using the Stata commands *tssmooth shwinters*. These algorithms use recent trends in a time-series to forecast future changes, placing declining value on older data, allowing recent trends to dominate predictions, and without including additional explanatory variables^11^. We made no attempt to use explanatory variables to inform modelling, given we examined trends across a broad range of causes.

## Findings

### Burden of disease in 2017

The proportion of current (2017) burden of disease attributable to each cause amongst 1-4 year olds are shown in Figure 1, with data for other age-groups shown in the Appendix (Figure A1 and table A3). Neonatal causes (diseases of prematurity) and congenital disorders dwarfed all other causes of DALYS across 0-24 year olds, with other the other main causes contributing to total burden being anxiety and depression, endocrine and immune disorders, and lower respiratory tract infections. Amongst infants, together 80% of burden is attributable to neonatal causes and congenital disorders, with lower proportions due to SIDS and lower respiratory tract infections, endocrine disorders. For 1-4 year olds, whilst neonatal and congenital causes continue to be responsible for 12.3% and 10.8% of total burden respectively, the largest cause was skin problems (22.5%), with other major causes being wheeze/asthma, common infections, unintentional injuries and diarrhoea. Of note, cancer and long-term conditions (asthma, epilepsy) begin to emerge as important causes in this age group. The distribution of causes for 5-9 year olds were similar to 1-4 year olds with skin still the largest cause of DALYS, although proportions due to neonatal and congenital disorders are lower than younger children and asthma, nutrition, neurodevelopmental problems and mental health issues emerge as substantial causes. By 10-14 years, mental health problems emerge as the largest cause of burden in both sexes (22.4% in males; 25.2% in females) with major other causes being skin, injuries, somatic symptoms (headache, pain) and long-term conditions. The predominance of mental health causes was also evident in 15-19 year olds (22.4% of males; 24.6%) and 20-24 year olds (18.5% males; 20.1% females), with the other largest causes being substance use, somatic symptoms and injuries.

**Figure 1.**
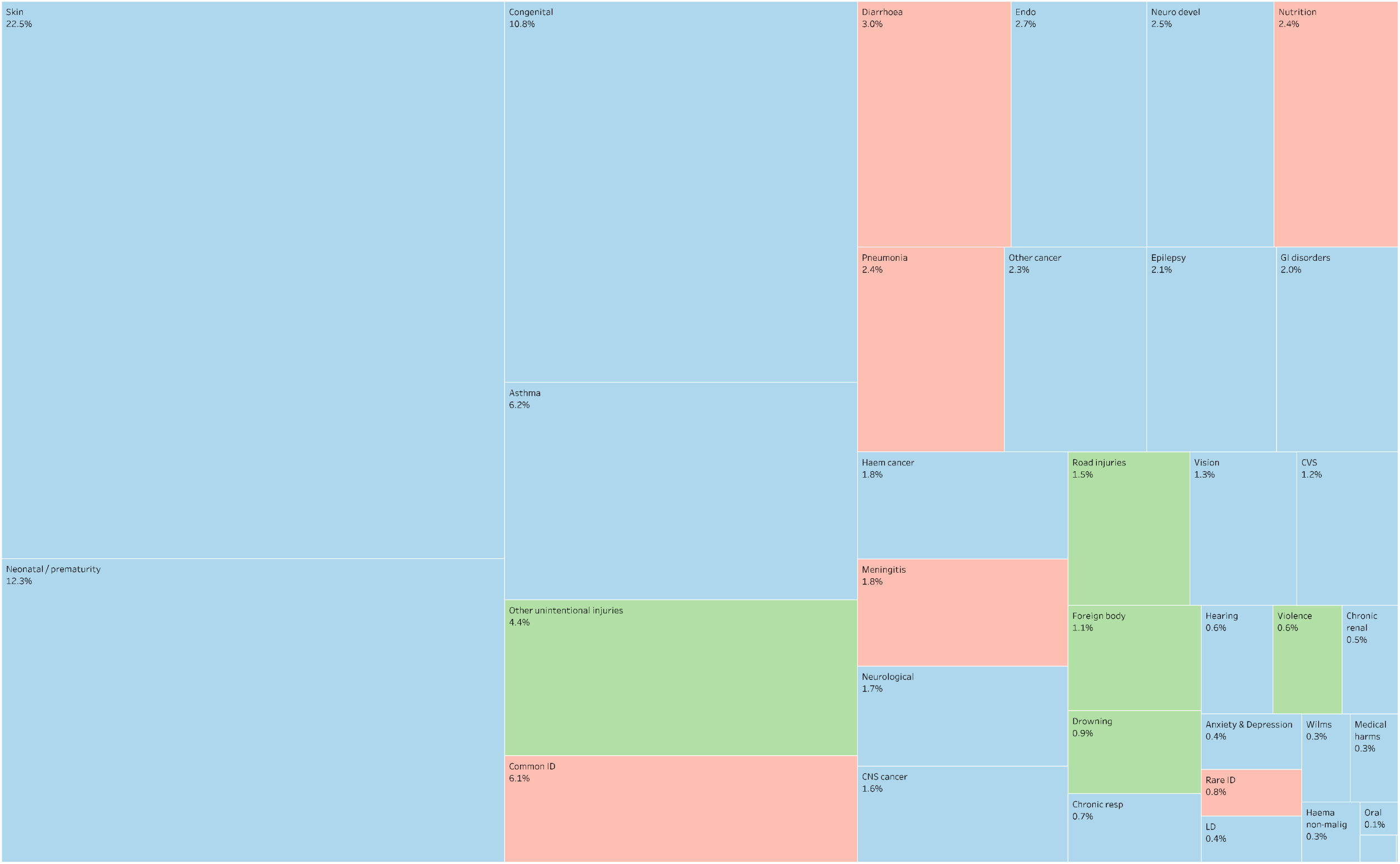
Treemap charts of proportions of total DALYs/100 000 population amongst 1-4 year olds for 2017 (communicable, nutritional & maternal conditions in red; neo-natal and non-communicable diseases in blue; injuries in green)

### Change in burden of disease in 1998 -2017

Trends in total DALYS per 100,000 population from 1998 to 2017 are shown for each age-group in Figure 2. Total DALYS were highest amongst neonates (largely reflecting the impact of infant mortality), and lowest amongst 1-9 year olds, rising with age amongst 10-24 year olds. Between 1998-2017, total DALYs fell in each age-group, with the most dramatic falls in infant age groups (fall of 43.8% amongst <7D, 37.4% amongst 7-28D, 48.8% in the post-neonatal period) and less amongst older age-groups (31.5% 1-4y; 19.6% 1-9y; 12.8% 10-14y; 15.5% 15-19y and 13.0% 20-24y). In each age-group, reductions plateaued after 2010/11.

Percent change in DALYs by cause over the last decade (2008 to 2017) is shown by age and sex in Figure 3, with detail shown in Appendix Table A4. Reductions in DALYS amongst neonates over the last decade have very largely comprised reductions in neonatal causes. Amongst post-neonatal infants, each of the main causes (neonatal causes, congenital, Sudden Infant Death Syndrome) fell 10-20%, with the only rise being in iron deficiency. In contrast, neonatal causes rose approximately 10% in each age group, likely reflecting the increased burden of neonatal survivorship. This burden was highest amongst 1-4 year olds (where neonatal causes represent 12% of total burden) but is evident through to 20-24 year olds (approximately 2.5-3% of burden).

**Figure 2.**
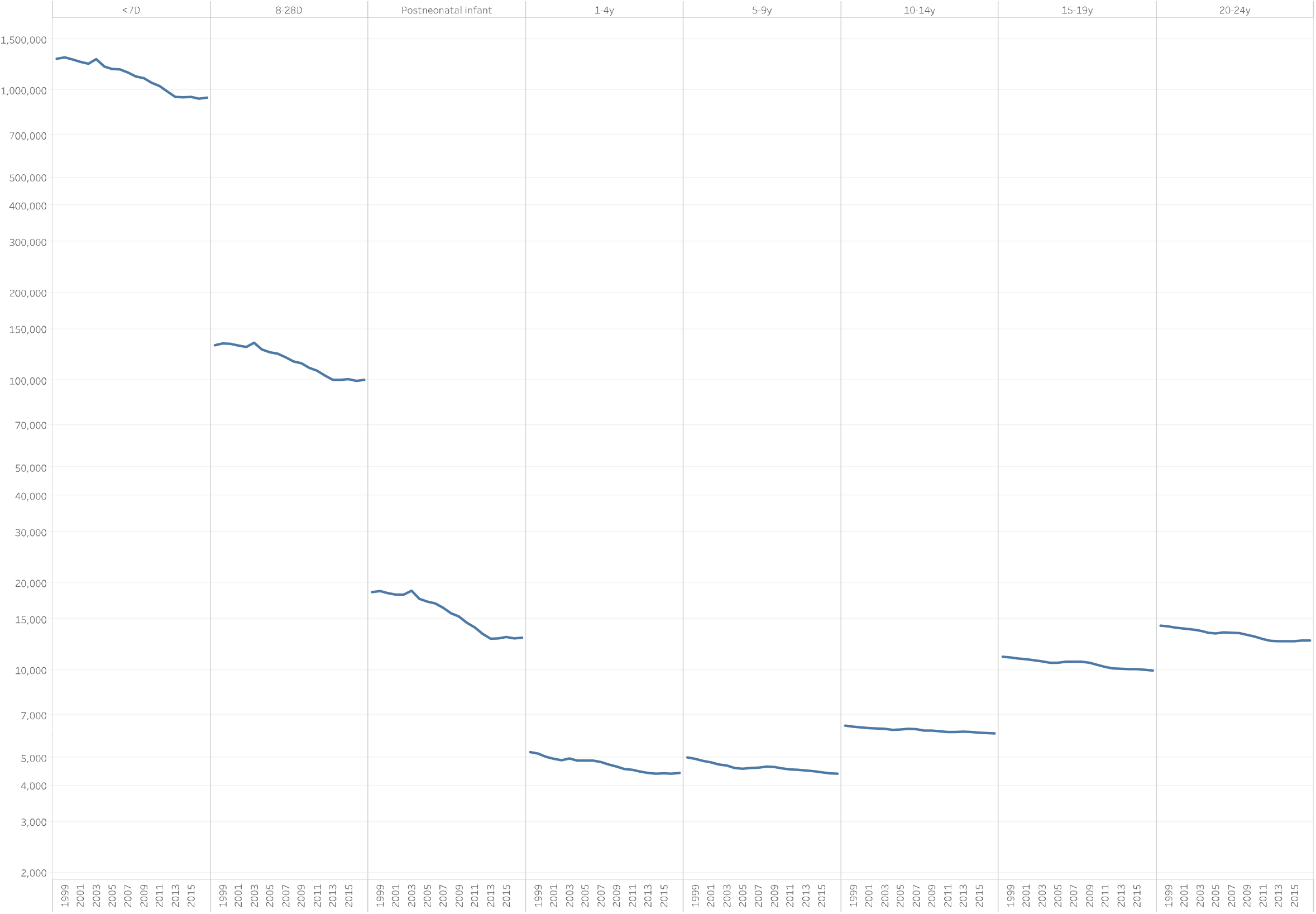
Trends in total DALYs per 100,000 by age-group in the UK, 1998 to 2017

**Figure 3.**
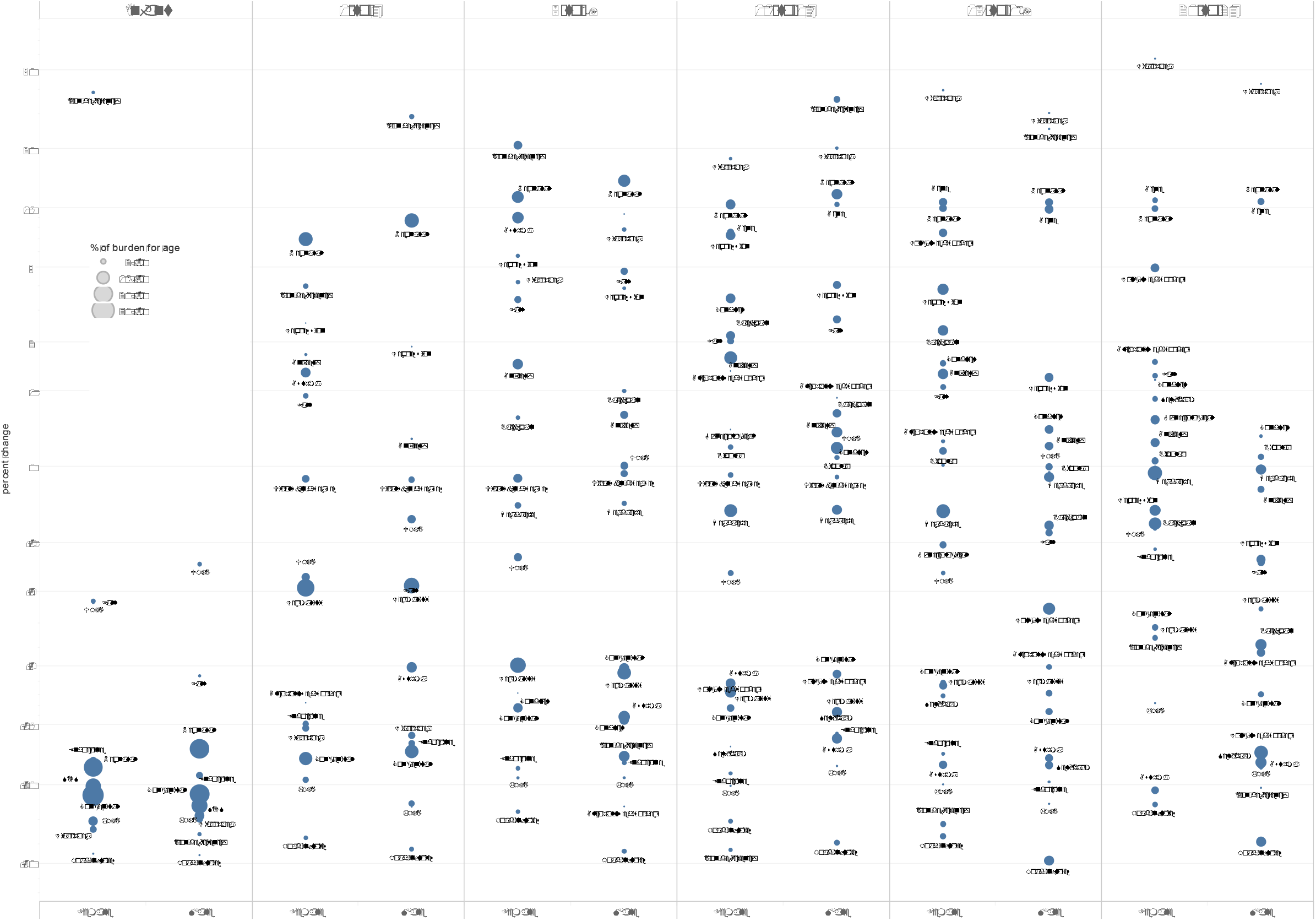
Proportional change in DALYS by cause from 2008 to 2017 by age

Amongst 1-4 year olds, rises were seen in neonatal causes, iron deficiency and mental health causes, with falls in all other causes, particularly for congenital causes. For 5-9 year olds, rises were seen in neonatal causes, fall injuries, mental health causes, iron deficiency and diarrhoea, with falls in causes such as skin, asthma, congenital causes and road injuries. Similar pattern was seen amongst 10-14 year olds, with additional rises in acne and back pain, whilst burden from asthma, skin, congenital causes and road injuries fell. Change was similar across both sexes amongst 15-19 year olds, with rises predominantly in mental health causes and alcohol and drug-use disorders, acne and neonatal causes, with notable falls in burden from road injuries, asthma and self-harm. Amongst 20-24 year olds, there were fewer rises than in 15-19 year olds although patterns amongst causes were similar.

### Forecasting future DALY burden

Forecast change in total DALYs from 2017 to 2040 are shown in Appendix Figure A2. Models suggest continued falls in total DALYS in all age-groups, continuing trends seen across 1990 to 2017, although these changes are only significant amongst infants and 15-19 year old males, with smaller reductions seen in other age-groups, reflecting recent plateaus in DALYs as noted above.

Forecast percent change by cause for the decades to 2030 and 2040 together with observed change for the decades to 2007 and 2017 are shown for 0-9 year olds in Table 1 and 10-24 year olds in Table 2. Reductions of ≥10% are shown in green, with increases ≥10% shown in red.

**Table 1.**
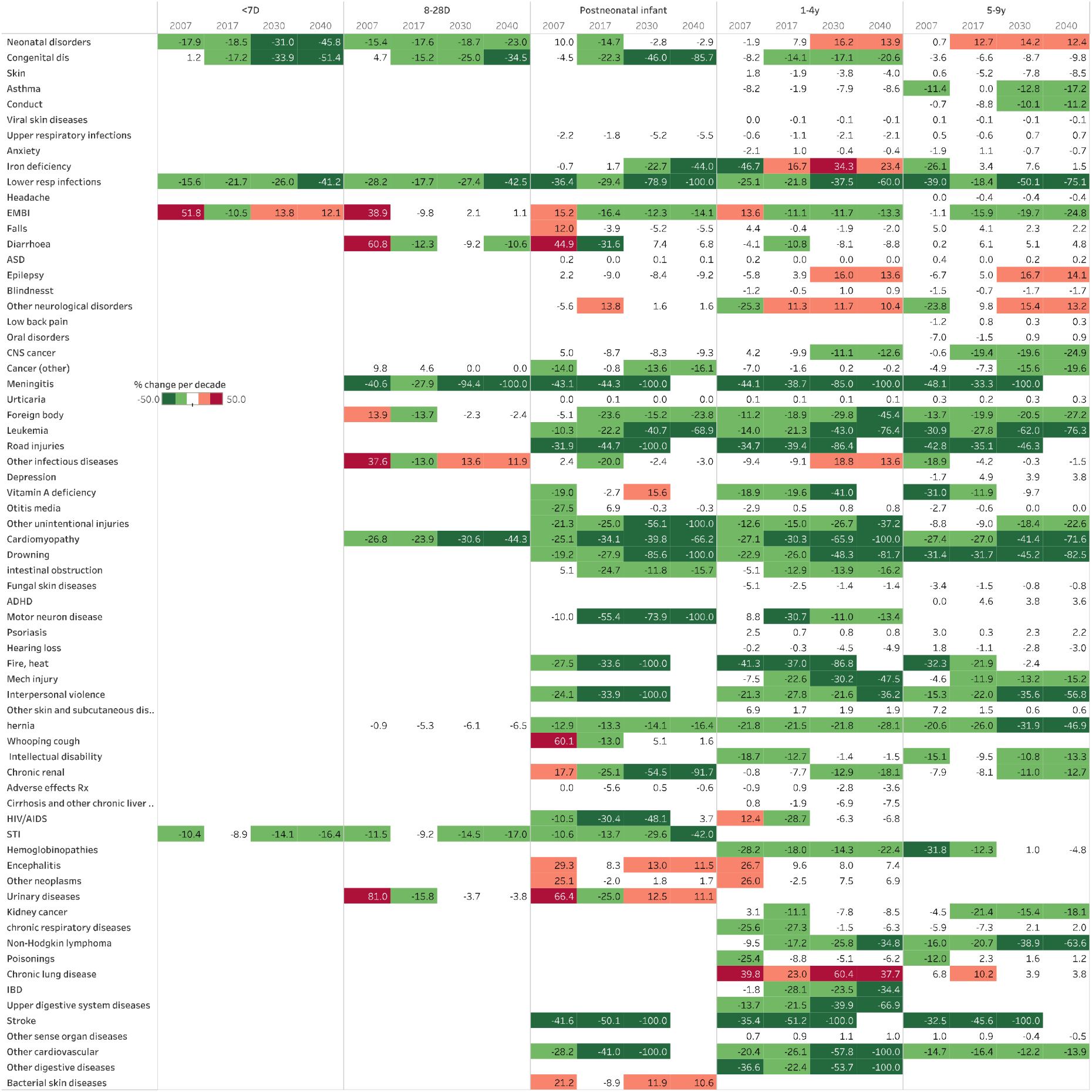
Percent change by cause for the decades 1998-2007 and 2008-2017 (observed) and forecast change for the decades to 2030 and 2040 for 0-9 year olds Note: Only major causes, i.e. those contributing 0.5% or more of total DALYS by age in 2017 are shown.

**Table 2.** Percent change by cause for the decades 1998-2007 and 2008-2017 (observed) and forecast change for the decades to 2030 and 2040 for 10-24 year olds (both sexes) Note: Only major causes, i.e. those contributing 0.5% or more of total DALYS by age in 2017 are shown.

|  | 10-14y |  |  |  | 15-19y |  |  |  | 20-24y |  |  |  |
| --- | --- | --- | --- | --- | --- | --- | --- | --- | --- | --- | --- | --- |
|  | 2007 | 2017 | 2030 | 2040 | 2007 | 2017 | 2030 | 2040 | 2007 | 2017 | 2030 | 2040 |
| Headache | -0.2 | -0.5 | -0.8 | -0.8 | -0.2 | -0.3 | -0.4 | -0.4 | 0.0 | 0.0 | -0.1 | -0.1 |
| Low back pain | -0.3 | 1.4 | 0.8 | 0.8 | -1.6 | 0.8 | -1.8 | -1.9 | -6.7 | -2.3 | -0.5 | -0.5 |
| Anxiety | -1.9 | 1.0 | -0.2 | -0.2 | -1.2 | 0.8 | 0.1 | 0.1 | 0.0 | 0.0 | 0.0 | 0.0 |
| Drug use | -2.8 | -5.7 | -10.2 | -11.3 | 1.7 | 2.5 | 2.3 | 1.5 | 7.0 | -4.3 | -2.5 | -2.6 |
| Depression | -3.9 | 5.7 | 7.7 | 7.1 | -5.5 | 2.6 | -0.7 | -0.8 | -6.0 | -0.9 | -1.8 | -1.8 |
| Asthma | -6.4 | -8.9 | -36.0 | -55.6 | -0.9 | -15.2 | -67.2 | -52.3 | 1.2 | -18.6 | -19.4 | -28.9 |
| Neonatal disorders | 1.7 | 11.2 | 10.8 | 9.7 | 2.3 | 10.5 | 9.7 | 8.8 | 3.0 | 10.5 | 9.3 | 8.5 |
| Gynecological |  |  |  |  | 3.3 | -1.0 | -1.9 | -1.9 | 2.1 | 0.6 | 1.6 | 1.6 |
| Skin | -1.1 | -7.6 | -10.0 | -11.2 | -0.1 | -6.6 | -9.8 | -10.9 | 0.6 | -2.8 | -4.1 | -4.3 |
| Conduct | 2.5 | 1.9 | -8.9 | -10.0 | 0.9 | 1.0 | -2.3 | -2.4 | 0.3 | 0.8 | 0.3 | 0.3 |
| Road injuries | -37.8 | -34.7 | -22.1 |  | -28.6 | -42.1 | -94.7 |  | -24.6 | -31.8 | -83.6 |  |
| Self-harm | -13.9 | -8.4 | -24.1 | -31.8 | -20.0 | -11.5 | -19.6 | -24.6 | -25.7 | -7.2 | -22.5 | -31.5 |
| Falls | 3.3 | 2.4 | 2.3 | 2.3 | 2.3 | 0.1 | 0.6 | 0.5 | 1.4 | 0.0 | 0.1 | 0.1 |
| Congenital dis | -3.9 | -6.8 | -4.2 | -4.5 | -5.5 | -7.3 | -8.8 | -9.7 | -3.2 | -5.0 | -6.5 | -7.0 |
| Acne | 17.0 | 9.1 | 5.9 | 5.4 | 16.8 | 10.4 | 4.3 | 4.1 | 15.1 | 11.0 | 9.1 | 8.3 |
| Bipolar disorder | -1.4 | 0.2 | 0.0 | 0.0 | -1.8 | 0.1 | -0.4 | -0.4 | -2.4 | 0.1 | -0.7 | -0.7 |
| Alcohol use disorders |  |  |  |  | 2.7 | -2.4 | -7.2 | -7.8 | 6.3 | -1.3 | -6.9 | -8.1 |
| Eating disorders | 9.2 | 4.2 | 5.3 | 5.0 | 7.9 | 3.8 | 3.2 | 3.0 | 6.5 | 1.8 | -0.4 | -0.6 |
| Epilepsy | -2.7 | -2.4 | 14.7 | 12.8 | 1.7 | -13.7 | -10.5 | -11.7 | 1.2 | -15.2 | -6.9 | -8.3 |
| Iron deficiency | 32.1 | -3.3 | -42.5 | 13.1 | -17.1 | -3.0 | -2.6 | -4.0 | -3.3 | -12.1 | -9.1 | -36.4 |
| Upper respiratory infections | 0.6 | -0.6 | 0.6 | 0.6 | 0.7 | -0.7 | 0.4 | 0.4 | 0.6 | -0.5 | 0.0 | 0.0 |
| Neck pain | -1.9 | 0.0 | 0.5 | 0.5 | -14.1 | 1.3 | 4.8 | 4.5 | -19.3 | 3.1 | 11.8 | 10.6 |
| ASD | 0.2 | 0.0 | 0.1 | 0.1 | 0.0 | 0.1 | 0.0 | 0.0 | 0.0 | 0.2 | 0.4 | 0.4 |
| Diabetes mellitus | 51.7 | -15.3 | -42.7 | 12.7 | 98.0 | -8.0 | 24.2 | 19.5 | 64.5 | 8.7 | 8.7 | 6.9 |
| Blindnesst | -1.6 | -1.0 | -1.6 | -1.6 | -1.8 | -1.3 | -2.0 | -2.1 | -2.0 | -1.7 | -2.2 | -2.2 |
| Other mental disorders |  |  |  |  | -0.2 | -0.1 | -0.1 | -0.1 | 0.1 | 0.0 | 0.0 | 0.0 |
| Oral disorders | -1.9 | -0.6 | -0.5 | -0.5 | -2.8 | -0.7 | -1.2 | -1.3 | -5.3 | -1.5 | 1.0 | 0.9 |
| Viral skin diseases | -0.1 | -0.1 | -0.1 | -0.1 | 0.0 | 0.0 | 0.1 | 0.1 | 0.1 | 0.0 | 0.0 | 0.0 |
| Psoriasis | 2.7 | 1.3 | 1.6 | 1.6 | 2.6 | 1.6 | 2.2 | 2.1 | 2.7 | 2.1 | 2.7 | 2.6 |
| Other musculoskeletal disorders |  |  |  |  | -2.3 | -17.9 | -15.8 | -19.0 | 0.3 | -6.8 | -3.6 | -3.8 |
| Hearing loss | 3.7 | 1.2 | -0.9 | -0.9 | 7.3 | 7.2 | -4.7 | -5.1 | 8.6 | 9.6 | -4.0 | -4.3 |
| Cancer (other) | 0.3 | -4.5 | -1.6 | -1.9 | -2.6 | -8.2 | -12.6 | -14.6 | -6.1 | -1.9 | -3.3 | -4.6 |
| EMBI | 0.5 | -14.2 | -17.3 | -21.7 | -4.3 | -16.6 | -10.4 | -11.7 | -7.6 | -8.8 | -15.6 | -19.4 |
| Other neurological disorders | -14.5 | 9.3 | 14.8 | 12.8 | -13.0 | -5.7 | -28.2 | -41.2 | -2.2 | -2.7 | -6.4 | -7.2 |
| Upper digestive system diseases |  |  |  |  | 0.8 | 0.7 | -2.4 | -2.5 | 0.1 | 0.3 | -0.6 | -0.6 |
| Other unintentional injuries | -9.1 | -6.9 | -13.2 | -15.4 | -11.4 | -8.0 | -12.2 | -14.0 | -11.1 | -5.9 | -6.8 | -7.3 |
| Interpersonal violence | -8.7 | -17.4 | -23.7 | -31.6 | -7.9 | -22.5 | -33.1 | -54.4 | -7.6 | -23.5 | -21.3 | -31.8 |
| Leukemia | -30.6 | -25.4 | -46.3 | -72.6 | -27.1 | -25.6 | -39.2 | -62.0 | -19.6 | -21.2 | -30.7 | -46.9 |
| Mech injury | -3.0 | -4.7 | -4.3 | -4.5 | -5.1 | -6.3 | -9.2 | -10.8 | -5.1 | -4.0 | -0.9 | -1.0 |
| CNS cancer | -2.7 | -13.4 | -17.5 | -21.2 | 5.1 | -12.3 | -5.5 | -6.2 | 0.6 | -5.1 | -4.8 | -5.1 |
| Schizophrenia |  |  |  |  | 0.7 | 3.4 | 1.9 | 1.9 | 0.8 | 7.0 | 9.1 | 8.3 |
| Urticaria | 0.1 | 0.2 | 0.1 | 0.1 | 0.0 | 0.0 | 0.0 | 0.0 | 0.1 | 0.0 | 0.1 | 0.1 |
| Foreign body | -13.3 | -18.3 | -17.0 | -25.9 | -30.8 | -24.9 | -86.4 | -100.0 | -28.8 | -21.9 | -31.5 | -46.0 |
| ADHD | -0.3 | 4.4 | 3.6 | 3.4 | -0.1 | 3.4 | 2.8 | 2.7 |  |  |  |  |
| Diarrhoea | 4.1 | 19.2 | 9.2 | 8.4 | 17.3 | 35.2 | 15.6 | 13.5 | 34.9 | 50.2 | 21.7 | 17.8 |
| Drowning | -27.0 | -21.1 | 5.1 | 4.9 | -18.2 | -29.6 | -58.6 | -100.0 | -26.1 | -19.6 | -45.9 | -85.0 |
| IBD | 39.1 | -2.2 | 1.2 | 0.1 | 46.5 | 2.5 | 1.6 | -2.3 | 42.8 | 5.3 | -4.7 | -6.2 |
| Other cardiovascular |  |  |  |  | -11.7 | -13.4 | -16.7 | -20.0 | -6.5 | -15.4 | -15.3 | -18.8 |
| chronic respiratory diseases | 11.0 | -0.2 | 3.7 | 3.5 |  |  |  |  |  |  |  |  |
| Stroke | -32.7 | -35.7 | -77.9 | -100.0 | -26.0 | -34.7 | -73.2 | -100.0 | -25.0 | -27.1 | -48.8 | -95.4 |
| Chronic lung disease | 3.7 | -3.1 | -7.9 | -10.2 | 4.3 | -15.9 | -31.3 | -54.9 | 5.5 | -19.2 | -53.5 | -66.6 |
| Cardiomyopathy | -30.9 | -25.7 | -50.7 | -100.0 | -16.4 | -31.3 | -40.6 | -68.3 | -13.6 | -18.6 | 4.9 | 1.9 |
| Fungal skin diseases | -1.3 | -0.5 | -0.3 | -0.3 | -0.8 | -0.3 | -0.1 | -0.1 | -0.5 | -0.2 | -0.1 | -0.1 |
| Other skin and subcutaneous dis.. | 7.0 | 1.9 | 0.6 | 0.6 | 6.8 | 2.0 | 0.7 | 0.7 | 6.9 | 2.2 | 2.5 | 2.4 |
| Other transport injuries | -12.4 | -12.7 | -10.6 | -11.9 | -3.9 | -22.7 | -50.4 | -100.0 | -14.3 | -15.1 | -33.4 | -50.1 |
| Otitis media | -4.6 | 0.0 | -0.9 | -1.0 | -5.1 | 0.3 | -1.9 | -1.9 |  |  |  |  |
| Lower resp infections | -38.7 | -18.1 | -42.1 | -74.0 | -39.6 | -22.3 | -24.6 | -35.1 | -42.5 | -11.9 | -23.6 | -36.7 |
| Poisonings | -19.1 | -6.7 | -4.3 | -5.1 | -37.3 | -15.4 | -10.1 | -11.2 | -39.3 | -20.6 | -0.2 | -0.2 |
| Intellectual disability | -15.1 | -9.5 | -10.3 | -12.7 | -11.3 | -2.7 | -2.7 | -2.7 |  |  |  |  |
| Hemoglobinopathies | 25.6 | -10.0 | 16.9 | 14.5 | 2.3 | -37.1 | -88.1 | -100.0 | -32.6 | -18.0 | -67.6 | -100.0 |
| Maternal disorders |  |  |  |  |  |  |  |  | -13.0 | -33.7 | -55.5 | -100.0 |
| Fire, heat | -22.6 | -10.4 | -5.7 |  | -24.0 | -10.5 | -36.3 |  | -26.1 | -14.0 | -36.7 |  |
| Rheumatoid arthritis | 24.1 | 3.2 | 0.7 | 0.7 | 23.7 | 6.3 | 2.7 | 2.6 | 22.2 | 9.4 | 5.0 | 4.8 |
| Meningitis | -49.2 | -30.3 | -97.3 | -100.0 | -53.6 | -35.0 | -84.8 | -100.0 |  |  |  |  |
| Other infectious diseases | 9.6 | 8.7 | 9.7 | 8.9 | -2.3 | -21.2 | -31.1 | -45.2 |  |  |  |  |
| Non-Hodgkin lymphoma | -16.1 | -19.7 | -30.2 | -43.3 | -18.6 | -25.3 | -26.0 | -35.1 | -24.3 | -24.0 | -27.4 | -37.7 |
| Chronic renal | -4.0 | -6.2 | -5.3 | -5.6 |  |  |  |  | -0.3 | -2.0 | -2.0 | -2.1 |

Forecasts for neonates predict continuation of major falls in neonatal and congenital causes and infectious diseases causes such as lower respiratory infections and meningitis over the next 20 years.

Amongst infants after the neonatal period, forecasts predict the contribution of neonatal causes will remain largely stable with large reductions predicted in burden from congenital disorders, infectious causes (lower respiratory, meningitis), injuries, cardiovascular disorders and leukaemia.

Amongst children 1-9 years, there are forecast rises in burden due to neonatal disorders, epilepsy and other neurological conditions and chronic lung disorders, although most of these rises are relatively small. In contrast, forecasts predict large declines in historic causes of burden including congenital causes, various cancers, injuries, asthma, infections and cardiovascular causes.

Amongst 10-24 year olds, forecasts predict that the large proportion of burden due to somatic symptoms, mental health problems and skin conditions will continue largely unchanged, with no conditions forecast to increase consistently across age-groups aside from diarrhoea. However, each older age-group is predicted to experience large falls in burden due to injuries including self-harm, various cancers and the majority of medical causes including asthma. These falls mean that the proportion of burden due to mental health problems and somatic symptoms will increase amongst 10-24 year olds over the next 20 years.

## Discussion

This is the first study to systematically examine recent trends in burden of disease amongst CYP in the UK and to use these data to forecast potential trends over the next two decades. We found that DALYS due to neonatal and congenital causes were the commonest cause of burden in the early life course from birth to age 24 years, likely reflecting both the high mortality rate of the early neonatal period and the high levels of sequelae of surviving extreme premature delivery. The predominant causes of burden of disease in younger children are common conditions readily recognised by paediatricians and general practitioners, including skin problems, wheeze and asthma, common infections, injuries and diarrhoea. In contrast, the predominant burden of disease in adolescents and young adults are mental health problems, somatic symptoms such as pain and headache, injuries and long-term conditions. Of these conditions affecting 10-24 year olds, only a minority have been historically the province of paediatricians.

We forecast future DALYS based upon trends over the past two decades. Our forecasts suggest that there will be continued falls in overall DALYS amongst infants over the next two decades, potentially reflecting continued gains in neonatal survival. However we forecast there will be few changes in the overall DALY burden in older age-groups, although there will be considerable shifts in the relative contribution of different causes. For younger children 1-9 years we forecast rises in burden due to the sequelae of neonatal survival including neurological conditions, epilepsy and chronic lung disease. These rises are also visible at a lower level amongst young people 10-24 years. Our forecasts suggest that there will be very large falls in many of the causes that have historically dominated disease in CYP, particularly congenital disorders, many infectious diseases, cancers and injuries. These declines likely represent falls in the prevalence of many infectious diseases and improvements in road safety but also marked improvements in survival from cancer and many congenital conditions.

The changes forecast here have implications for the skills and training of health professionals working with CYP over the next 20 years. However care is needed as changes in overall DALYS may represent a change in the balance of YLLs and YLDs for a condition, as increased survival from many childhood conditions (resulting in reduced YLLs) will inevitably increase morbidity (i.e. YLDs) in survivors. For example, largely stable forecast DALYS in conditions with little change in forecast burden, such as chronic renal disease, may reflect the inherent rise in morbidity (YLDs) that occurs with increased survival (i.e. reduced YLLs). In contrast, overall falls in DALYS amongst neonatal causes amongst infants and in cancer in older CYP likely reflect very large improvements in survival that are greater than the rises in morbidity in survivors. Therefore reductions in DALYs do not necessarily indicate a reduced need for workforce or skills related to particular causes, and indeed may be a marker of the impact and quality of current workforce inputs.

Forecast increases in DALYS from particular causes are, however, likely to indicate a need for additional workforce or a change in training focus. For paediatricians, there will be a need for broader training that includes increased numbers with skills in dealing with mental health problems, broader adolescent health issues as well as the consequences of neonatal survival, such as neuro-disability and epilepsy. This particularly important given the shift towards integrated 0 to 24 year services in England10 and likely similar moves in other countries.

Our data are subject to a number of limitations. DALY estimates take no account of co-morbidity, e.g. coexistent eczema and asthma, and co-morbidities may be increasing in the CYP population, e.g. children with exceptional needs. Data used were from the 2017 GBD study and therefore take no account of burden associated with COVID-19 nor of potential future conditions (e.g. long COVID). The forecasting algorithms use historical trend data over the past 20 years to project future trends. The priority given within the algorithm to more recent years provides an advantage over regression-based models which average change over the historical period. However it is important to note that these forecasts are merely projections of continuations of historical trends, and take no account of changes in the broader structural determinants of health or in the health system. Our findings of very high auto-correlation in the historic DALY series supports the contention that historic trends are an appropriate guide to future trends. We restricted these analyses to DALYS, which provide an integrative measure of both mortality and morbidity, to reduce complexity and to limit the amount of forecasting undertaken. However these estimates of total burden do not allow more detailed estimates of potential future change in hospital inpatient activity (more aligned with YLL burden) compared with ambulatory and community care (more aligned with YLDs). The level 3 GBD categorisation we used for causes does not align highly with work currently undertaken by paediatricians. Whilst we recategorized some causes to improve this alignment, other areas of work undertaken by paediatricians (e.g. child protection work; many aspects of community paediatrics) are not well represented in the cause hierarchy. Our analyses were restricted to the UK however there are likely to be differing trends in each UK country and regionally within England.

## Conclusions

Burden of health and illhealth in CYP is likely to change considerably over the next two decades if recent trends continue. The pattern of health and disease facing child health professionals in 2040 is likely to consist of higher proportions of mental health, other adolescent health issues, neurodisability and long-term conditions than currently, and this must be reflected in changes in training requirements for paediatricians and other child health professionals. More detailed work is needed to understand the balance of change in mortality (YLLs) and morbidity (YLDs) for this future burden, differences across UK countries, the impact of deprivation and ethnicity for changing patterns of burden. The impact of the COVID-19 pandemic on forecast changes is unclear, although it is most likely that the pandemic may accelerate predicted rises in burden due to mental health problems and slow predicted falls in other causes. Similarly, further work is needed to understand the impact of climate change and changes in child poverty on health burden for CYP over the next two decades.

## Data Availability

Data are publicly available from http://www.healthdata.org

