## Appendix for "Change in burden of disease in UK children and young people (0-24 years) over the past 20 years and estimation of potential burden in 2040: analysis using Global Burden of Disease (GBD) data"

Appendix GBD burden v1

#### Table A1. Mapping of GBD Level 3 causes to cause groups for these analyses

Notes: These modifications were made by expert consensus within the Paediatrics 2040 working group. For example all non-epilepsy neurological causes were grouped together, as many such causes were very rare in CYP (Alzheimer disease, motor-neuron disease, Parkinson’s) and the majority of paediatric neurological conditions were already grouped at Level 3 in Other neurological disorders). Causes with low levels of DALYs were grouped together where possible whilst more common causes were not. We also minimally modified some cause names to render them more comprehensible. ID=infectious diseases

| **Cause group** | **GBD Level 3 Cause** | Level 3 cause ID |
| --- | --- | --- |
| **Adverse effects of medical treatment** | Adverse effects of medical treatment | 708 |
| **Arthritis** | Gout | 632 |
|  | Osteoarthritis | 628 |
|  | Other musculoskeletal disorders | 639 |
|  | Rheumatoid arthritis | 627 |
| **Asthma** | Asthma | 515 |
| **Penumonia** | Lower respiratory infections | 322 |
| **Chronic resp** | Chronic obstructive pulmonary disease | 509 |
|  | Interstitial lung disease and pulmonary sarcoidosis | 516 |
|  | Other chronic respiratory diseases | 520 |
|  | Pneumoconiosis | 510 |
| **Cardiovascular** | Aortic aneurysm | 501 |
|  | Atrial fibrillation and flutter | 500 |
|  | Cardiomyopathy and myocarditis | 499 |
|  | Endocarditis | 503 |
|  | Hypertensive heart disease | 498 |
|  | Ischemic heart disease | 493 |
|  | Non-rheumatic valvular heart disease | 504 |
|  | Other cardiovascular and circulatory diseases | 507 |
|  | Peripheral artery disease | 502 |
|  | Rheumatic heart disease | 492 |
|  | Stroke | 494 |
| **Chronic renal** | Acute glomerulonephritis | 588 |
|  | Chronic kidney disease | 589 |
|  | Urinary diseases and male infertility | 594 |
| **Haem cancer** | Hodgkin lymphoma | 484 |
|  | Leukemia | 487 |
|  | Multiple myeloma | 486 |
|  | Non-Hodgkin lymphoma | 485 |
| **CNS cancer** | Brain and nervous system cancer | 477 |
| **Other cancer** | Bladder cancer | 474 |
|  | Breast cancer | 429 |
|  | Cervical cancer | 432 |
|  | Colon and rectum cancer | 441 |
|  | Esophageal cancer | 411 |
|  | Gallbladder and biliary tract cancer | 453 |
|  | Kidney cancer | 471 |
|  | Larynx cancer | 423 |
|  | Lip and oral cavity cancer | 444 |
|  | Liver cancer | 417 |
|  | Malignant skin melanoma | 459 |
|  | Mesothelioma | 483 |
|  | Nasopharynx cancer | 447 |
|  | Non-melanoma skin cancer | 462 |
|  | Other malignant neoplasms | 489 |
|  | Other neoplasms | 490 |
|  | Other pharynx cancer | 450 |
|  | Ovarian cancer | 465 |
|  | Pancreatic cancer | 456 |
|  | Prostate cancer | 438 |
|  | Stomach cancer | 414 |
|  | Testicular cancer | 468 |
|  | Thyroid cancer | 480 |
|  | Tracheal, bronchus, and lung cancer | 426 |
|  | Uterine cancer | 435 |
| **Haema non-malig** | Hemoglobinopathies and hemolytic anemias | 613 |
| **Congenital** | Congenital birth defects | 641 |
| **Diabetes** | Diabetes mellitus | 587 |
| **Endo** | Endocrine, metabolic, blood, and immune disorders | 619 |
| **GI disorders** | Appendicitis | 529 |
|  | Cirrhosis and other chronic liver diseases | 521 |
|  | Gallbladder and biliary diseases | 534 |
|  | Inflammatory bowel disease | 532 |
|  | Inguinal, femoral, and abdominal hernia | 531 |
|  | Other digestive diseases | 541 |
|  | Pancreatitis | 535 |
|  | Paralytic ileus and intestinal obstruction | 530 |
|  | Upper digestive system diseases | 992 |
|  | Vascular intestinal disorders | 533 |
| **Diarrhoea** | Diarrheal diseases | 302 |
| **Neurological** | Alzheimer's disease and other dementias | 543 |
|  | Motor neuron disease | 554 |
|  | Multiple sclerosis | 546 |
|  | Other neurological disorders | 557 |
|  | Parkinson's disease | 544 |
|  | Schizophrenia | 559 |
| **Epilepsy** | Epilepsy | 545 |
| **Foreign body** | Foreign body | 712 |
| **Gynaecological** | Gynecological diseases | 603 |
| **Headache** | Headache disorders | 972 |
| **Hearing** | Age-related and other hearing loss | 674 |
|  | Other sense organ diseases | 679 |
| **Common ID** | Bacterial skin diseases | 980 |
|  | Measles | 341 |
|  | Otitis media | 329 |
|  | Tuberculosis | 297 |
|  | Upper respiratory infections | 328 |
|  | Varicella and herpes zoster | 342 |
|  | Whooping cough | 339 |
| **Rare ID** | Acute hepatitis | 400 |
|  | African trypanosomiasis | 350 |
|  | Chagas disease | 346 |
|  | Cystic echinococcosis | 353 |
|  | Cysticercosis | 352 |
|  | Dengue | 357 |
|  | Diphtheria | 338 |
|  | Ebola | 843 |
|  | Food-borne trematodiases | 364 |
|  | Guinea worm disease | 936 |
|  | Intestinal nematode infections | 360 |
|  | Invasive Non-typhoidal Salmonella (iNTS) | 959 |
|  | Leishmaniasis | 347 |
|  | Leprosy | 405 |
|  | Lymphatic filariasis | 354 |
|  | Malaria | 345 |
|  | Onchocerciasis | 355 |
|  | Other intestinal infectious diseases | 321 |
|  | Other neglected tropical diseases | 365 |
|  | Other unspecified infectious diseases | 408 |
|  | Rabies | 359 |
|  | Schistosomiasis | 351 |
|  | Tetanus | 340 |
|  | Trachoma | 356 |
|  | Typhoid and paratyphoid | 958 |
|  | Yellow fever | 358 |
|  | Zika virus | 935 |
| **HIV/AIDS** | HIV/AIDS | 298 |
| **Meningitis** | Encephalitis | 337 |
|  | Meningitis | 332 |
| **Maternal** | Maternal disorders | 366 |
| **Neonatal / prematurity** | Neonatal disorders | 380 |
| **Neurodevelopmental** | Attention-deficit/hyperactivity disorder | 578 |
|  | Autism spectrum disorders | 575 |
|  | Other mental disorders | 585 |
| **Nutrition** | Dietary iron deficiency | 390 |
|  | Iodine deficiency | 388 |
|  | Other nutritional deficiencies | 391 |
|  | Protein-energy malnutrition | 387 |
|  | Vitamin A deficiency | 389 |
| **Oral** | Oral disorders | 680 |
| **Road injuries** | Other transport injuries | 695 |
|  | Road injuries | 689 |
| **Other unintentional injuries** | Animal contact | 709 |
|  | Environmental heat and cold exposure | 842 |
|  | Exposure to mechanical forces | 704 |
|  | Falls | 697 |
|  | Fire, heat, and hot substances | 699 |
|  | Other unintentional injuries | 716 |
|  | Poisonings | 700 |
| **Drowning** | Drowning | 698 |
| **Pain** | Low back pain | 630 |
|  | Neck pain | 631 |
| **SIDS** | Sudden infant death syndrome | 686 |
| **Acne** | Acne vulgaris | 661 |
| **Skin** | Alopecia areata | 662 |
|  | Decubitus ulcer | 665 |
|  | Dermatitis | 654 |
|  | Fungal skin diseases | 659 |
|  | Other skin and subcutaneous diseases | 668 |
|  | Pruritus | 663 |
|  | Psoriasis | 655 |
|  | Scabies | 658 |
|  | Urticaria | 664 |
|  | Viral skin diseases | 660 |
| **STD** | Sexually transmitted infections excluding HIV | 393 |
| **Vision** | Blindness and vision impairment | 981 |
| **Anxiety & Depression** | Anxiety disorders | 571 |
|  | Bipolar disorder | 570 |
|  | Depressive disorders | 567 |
| **Conduct** | Conduct disorder | 579 |
| **Self harm** | Self-harm | 718 |
| **Eating disorders** | Eating disorders | 572 |
| **Subs use** | Alcohol use disorders | 560 |
|  | Drug use disorders | 561 |
| **Violence** | Conflict and terrorism | 945 |
|  | Executions and police conflict | 854 |
|  | Exposure to forces of nature | 729 |
|  | Interpersonal violence | 724 |
| **LD** | Idiopathic developmental intellectual disability | 582 |

### Table A2. Correlation matrix of DALY values in 2017 with lagged values for the previous decade for the top 30 causes of DALYS for each age-group

|  | **Early Neonatal** |  | **Late Neonatal** | | **Post-neonatal infant** | | **1-4y** | | **5-9y** | | **10-14y** | | **15-19y** | | **20-24y** | |
| --- | --- | --- | --- | --- | --- | --- | --- | --- | --- | --- | --- | --- | --- | --- | --- | --- |
|  | *Coefficient* | *p* | *Coefficient* | *p* | *Coefficient* | *p* | *Coefficient* | *p* | *Coefficient* | *p* | *Coefficient* | *p* | *Coefficient* | *p* | *Coefficient* | *p* |
| **2016** | 1 | 8.50E-104 | 0.9999999 | 3.88E-93 | 0.9999989 | 1.58E-80 | 0.9999957 | 1.93E-72 | 0.9999968 | 2.49E-74 | 0.9999962 | 2.81E-73 | 0.9999956 | 2.72E-72 | 0.9999979 | 9.47E-77 |
| **2015** | 0.9999999 | 7.00E-96 | 0.9999992 | 6.04E-83 | 0.9999929 | 1.87E-69 | 0.9999883 | 2.11E-66 | 0.9999887 | 1.41E-66 | 0.999987 | 9.42E-66 | 0.9999864 | 1.90E-65 | 0.9999916 | 2.01E-68 |
| **2014** | 0.9999995 | 1.19E-85 | 0.9999988 | 2.43E-80 | 0.999991 | 5.95E-68 | 0.9999771 | 2.70E-62 | 0.9999737 | 1.83E-61 | 0.9999757 | 6.17E-62 | 0.999975 | 9.15E-62 | 0.9999819 | 1.00E-63 |
| **2013** | 0.9999782 | 1.38E-62 | 0.9999673 | 3.98E-60 | 0.9999446 | 6.21E-57 | 0.9999668 | 4.86E-60 | 0.9999604 | 5.77E-59 | 0.9999642 | 1.40E-59 | 0.9999617 | 3.59E-59 | 0.999972 | 4.53E-61 |
| **2012** | 0.9999962 | 2.94E-73 | 0.9999945 | 5.43E-71 | 0.9999725 | 3.50E-61 | 0.999946 | 4.43E-57 | 0.9999434 | 8.50E-57 | 0.9999534 | 5.53E-58 | 0.9999463 | 4.08E-57 | 0.9999663 | 5.99E-60 |
| **2011** | 0.9999964 | 1.24E-73 | 0.9999872 | 7.38E-66 | 0.9999638 | 1.62E-59 | 0.9999141 | 2.92E-54 | 0.9999293 | 1.92E-55 | 0.999944 | 7.26E-57 | 0.9999177 | 1.60E-54 | 0.9999636 | 1.78E-59 |
| **2010** | 0.9999899 | 2.66E-67 | 0.999971 | 7.30E-61 | 0.9999431 | 9.08E-57 | 0.9998881 | 1.18E-52 | 0.9999082 | 7.37E-54 | 0.9999345 | 6.48E-56 | 0.9998415 | 1.55E-50 | 0.9999287 | 2.14E-55 |
| **2009** | 0.9999997 | 2.38E-88 | 0.9999856 | 4.09E-65 | 0.9998959 | 4.29E-53 | 0.9998209 | 8.56E-50 | 0.9998752 | 5.46E-52 | 0.9999247 | 4.63E-55 | 0.9997052 | 9.14E-47 | 0.9998757 | 5.15E-52 |
| **2008** | 0.9999995 | 4.68E-85 | 0.9999866 | 1.42E-65 | 0.9998207 | 8.67E-50 | 0.9997298 | 2.70E-47 | 0.9998452 | 1.11E-50 | 0.9999127 | 3.65E-54 | 0.9995305 | 6.18E-44 | 0.9997939 | 6.12E-49 |
| **2007** | 0.9999949 | 1.77E-71 | 0.999977 | 2.79E-62 | 0.9997095 | 7.43E-47 | 0.9996223 | 2.94E-45 | 0.9998375 | 2.19E-50 | 0.9998666 | 1.39E-51 | 0.9994063 | 1.65E-42 | 0.999744 | 1.27E-47 |

Table shows average correlations of 2017 DALY values with lagged values for previous decade for top 30 causes of DALYS by age

#### Figure A1. Treemap charts of proportions of total DALYs/100 000 population by age-group for 2017

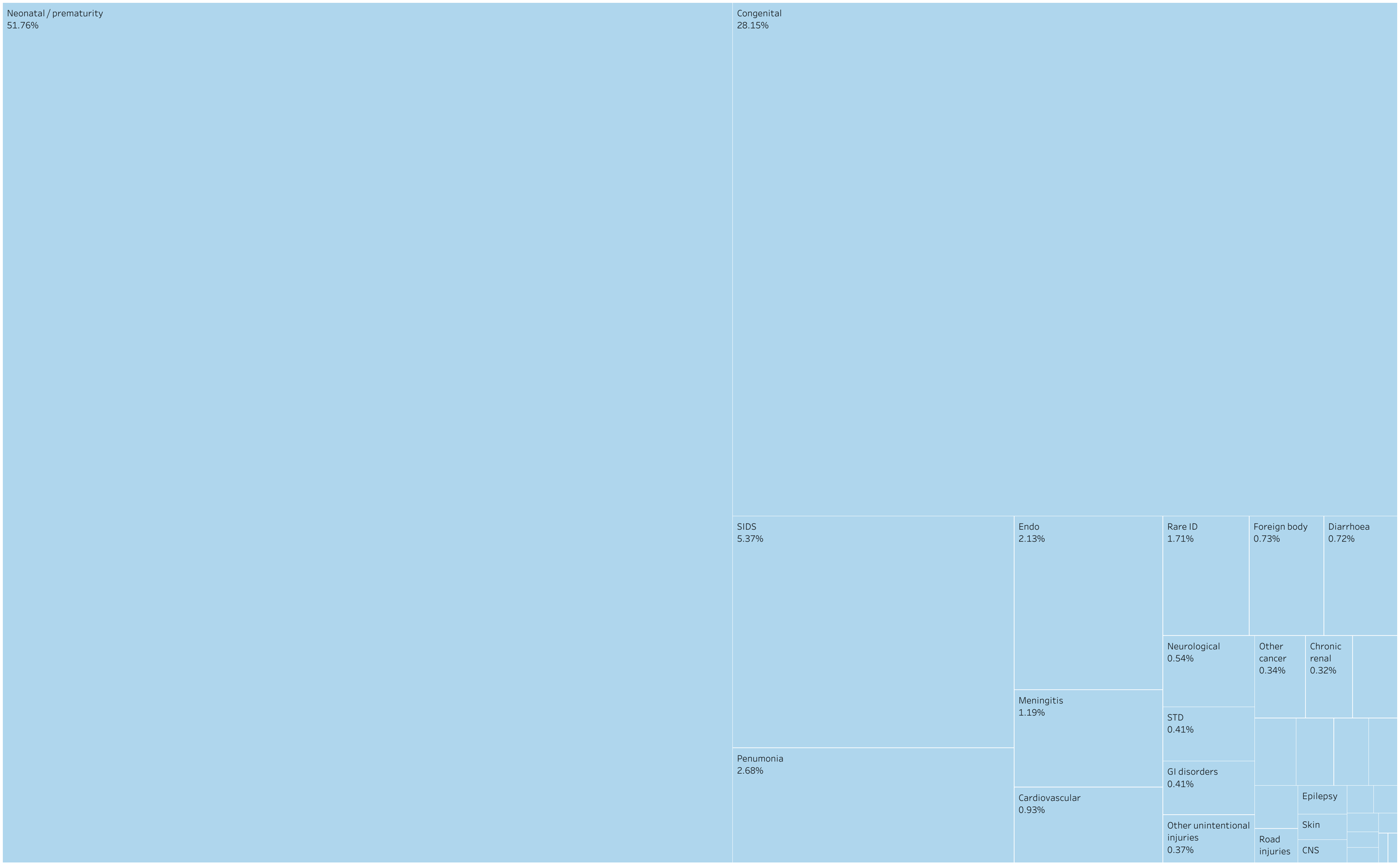

#### Infant

#### 5-9 years

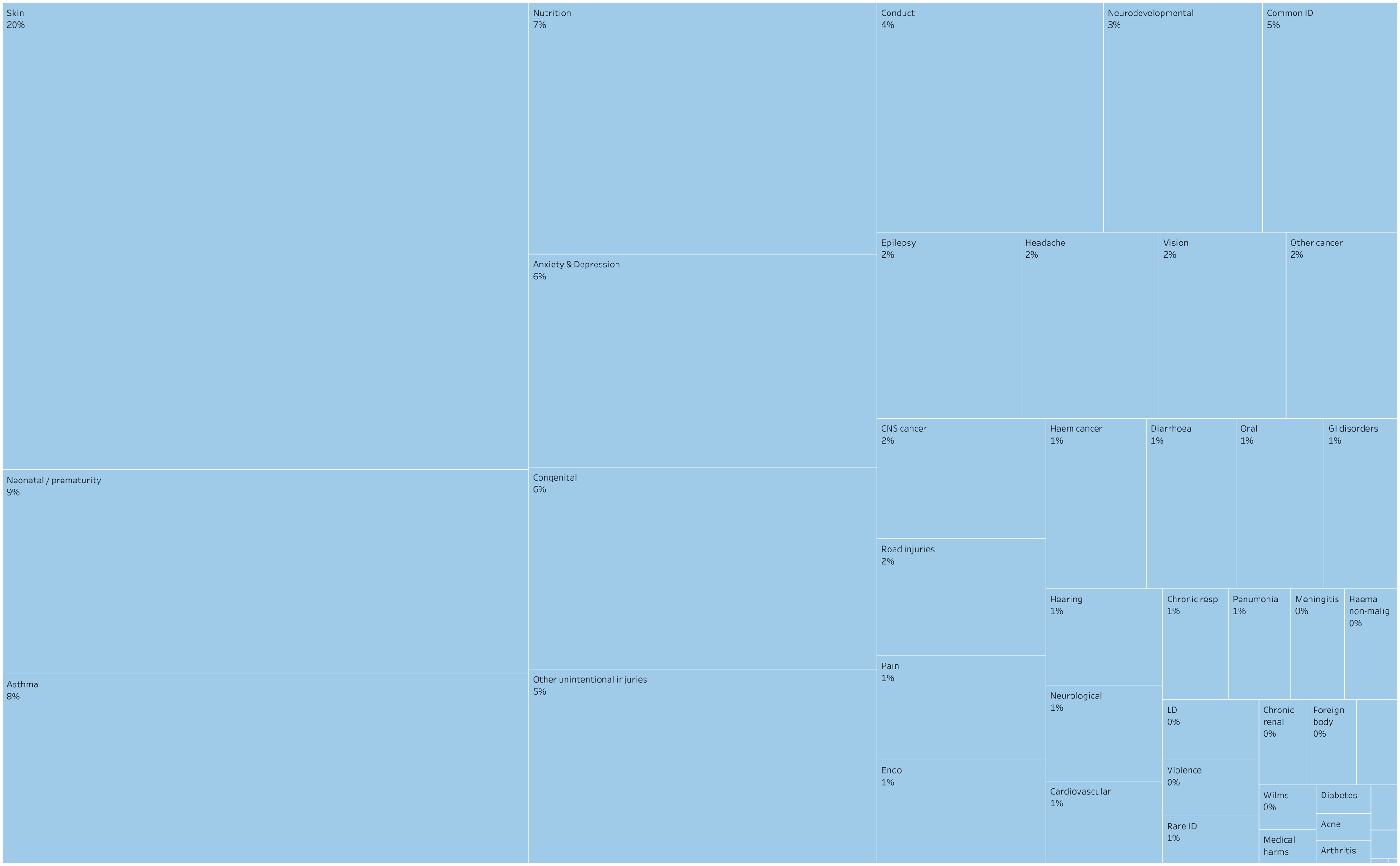

10-14 years

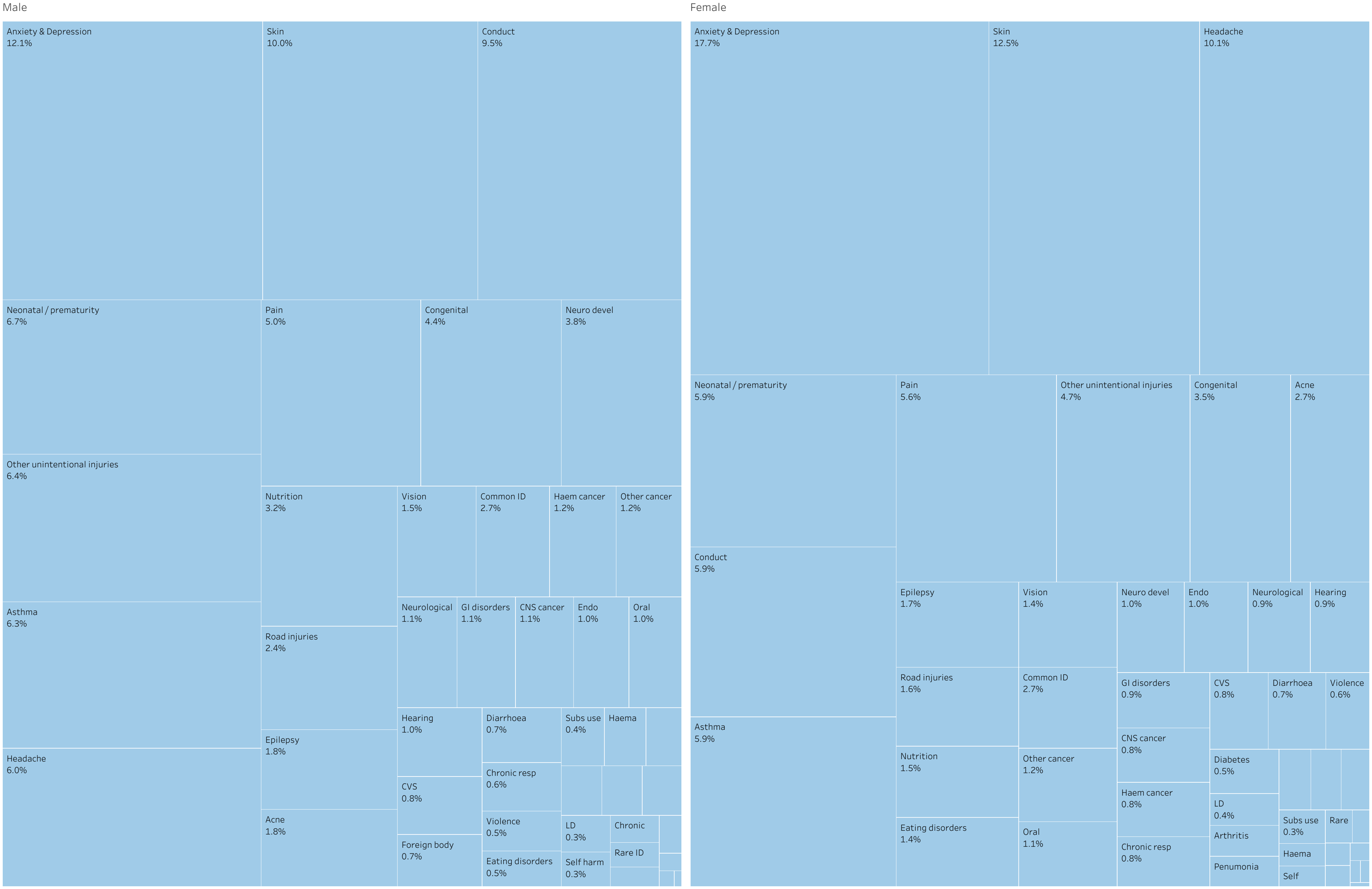

15-19 years

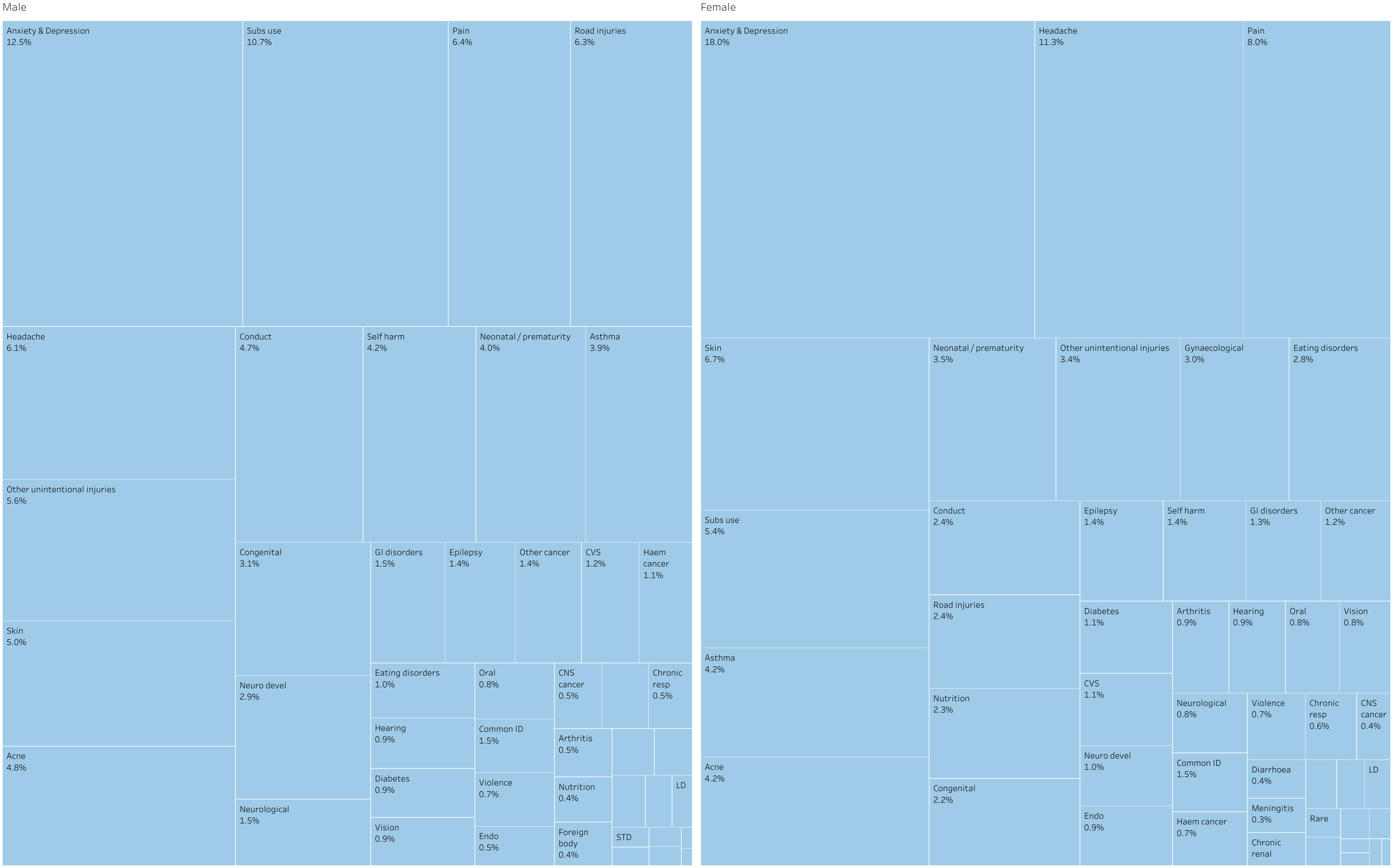

20-24 years

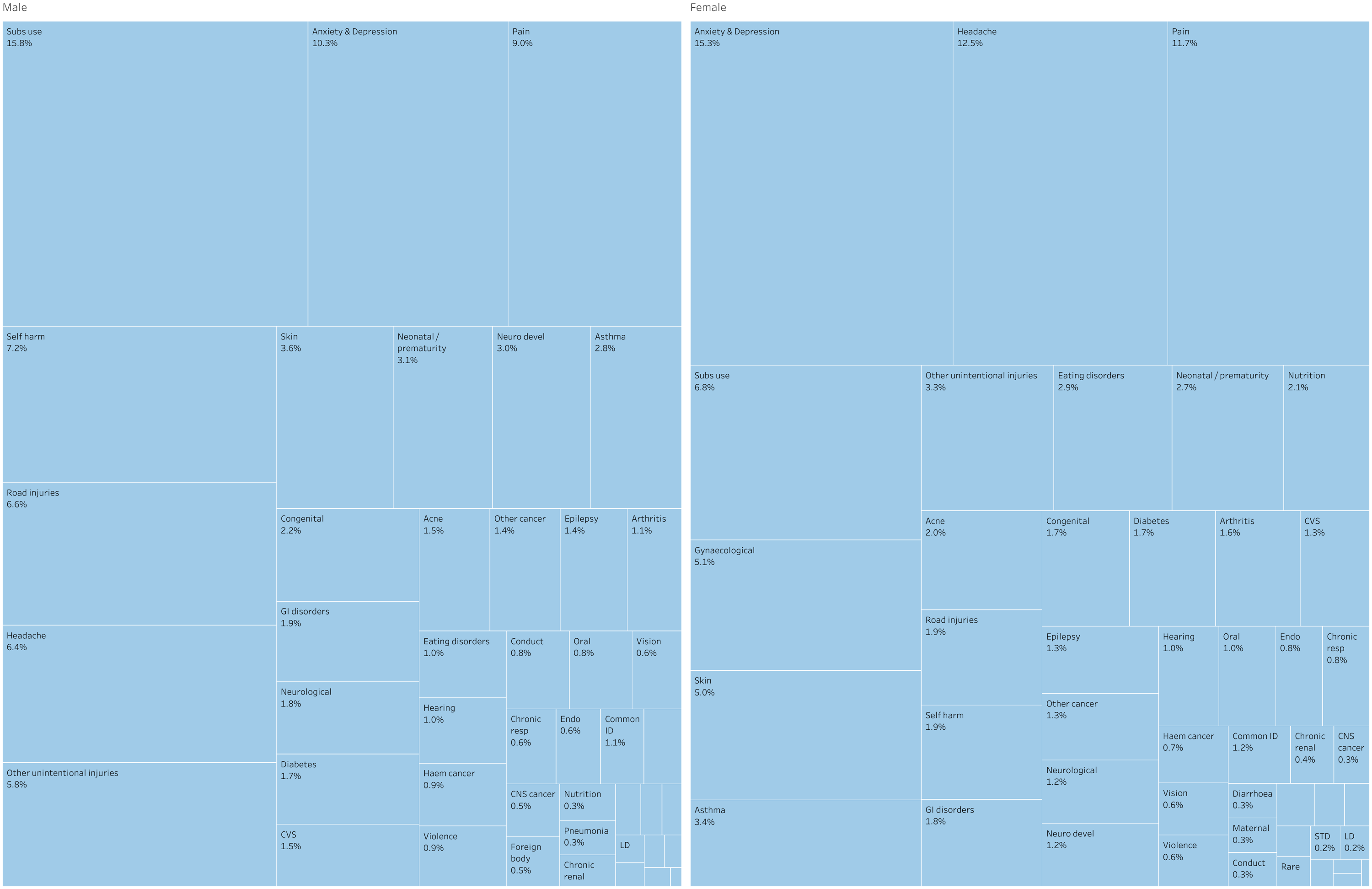

#### Table A3. DALYS per 100,000 by age, sex and cause in 2017

|  | **Early Neonatal** | **Late Neonatal** | **Post Neonatal** | **1 to 4** | **5 to 9** | **10 to 14** | | **15 to 19** | | **20 to 24** | | **Grand Total** |
| --- | --- | --- | --- | --- | --- | --- | --- | --- | --- | --- | --- | --- |
|  | **Both** | **Both** | **Both** | **Both** | **Both** | **Female** | **Male** | **Female** | **Male** | **Female** | **Male** |  |
| Acne | 0 | 0 | 0 | 0 | 5 | 170 | 104 | 436 | 460 | 261 | 183 | 1,620 |
| Anxiety & Depression | 0 | 0 | 0 | 20 | 265 | 1,100 | 720 | 1,855 | 1,210 | 1,973 | 1,282 | 8,426 |
| Arthritis | 0 | 0 | 0 | 0 | 4 | 22 | 10 | 91 | 46 | 213 | 140 | 526 |
| Asthma | 0 | 0 | 0 | 273 | 356 | 364 | 377 | 437 | 380 | 437 | 350 | 2,974 |
| Chronic renal | 559 | 232 | 134 | 22 | 15 | 20 | 13 | 34 | 23 | 54 | 37 | 1,143 |
| Chronic resp | 0 | 0 | 0 | 33 | 26 | 48 | 38 | 60 | 48 | 102 | 78 | 432 |
| CNS cancer | 317 | 56 | 52 | 72 | 73 | 52 | 64 | 40 | 52 | 45 | 59 | 881 |
| Common ID | 114 | 186 | 332 | 269 | 221 | 167 | 162 | 154 | 141 | 155 | 137 | 2,037 |
| Conduct | 0 | 0 | 0 | 0 | 186 | 365 | 564 | 248 | 452 | 36 | 103 | 1,954 |
| Congenital | 225,000 | 28,690 | 3,327 | 479 | 252 | 218 | 261 | 230 | 297 | 219 | 278 | 259,251 |
| CVS | 2,077 | 853 | 206 | 55 | 34 | 47 | 49 | 117 | 115 | 174 | 187 | 3,914 |
| Diabetes | 60 | 58 | 4 | 4 | 6 | 32 | 20 | 117 | 84 | 216 | 210 | 811 |
| Diarrhoea | 569 | 455 | 367 | 134 | 55 | 46 | 43 | 39 | 32 | 37 | 27 | 1,805 |
| Drowning | 101 | 19 | 27 | 39 | 13 | 6 | 20 | 7 | 50 | 9 | 60 | 351 |
| Eating disorders | 0 | 0 | 0 | 0 | 3 | 89 | 28 | 289 | 94 | 374 | 122 | 999 |
| Endocrinology and immune | 6,576 | 2,050 | 374 | 119 | 62 | 60 | 61 | 96 | 52 | 102 | 71 | 9,624 |
| Epilepsy | 40 | 40 | 92 | 95 | 95 | 108 | 108 | 146 | 140 | 170 | 174 | 1,209 |
| Foreign body | 604 | 551 | 283 | 50 | 14 | 18 | 44 | 17 | 42 | 24 | 56 | 1,704 |
| GI disorders | 395 | 283 | 180 | 89 | 45 | 53 | 65 | 131 | 147 | 231 | 241 | 1,859 |
| Gynaecological | 0 | 0 | 0 | 0 | 0 | 2 | 0 | 311 | 0 | 659 | 0 | 972 |
| Haem cancer | 300 | 116 | 66 | 80 | 61 | 52 | 73 | 71 | 105 | 87 | 115 | 1,128 |
| Haematology non-malignant | 132 | 48 | 21 | 13 | 21 | 11 | 24 | 24 | 10 | 28 | 10 | 341 |
| Headache | 0 | 0 | 0 | 0 | 92 | 626 | 357 | 1,160 | 588 | 1,614 | 793 | 5,228 |
| Hearing | 27 | 27 | 19 | 28 | 40 | 56 | 58 | 91 | 87 | 131 | 121 | 684 |
| HIV/AIDS | 0 | 0 | 56 | 9 | 4 | 4 | 2 | 6 | 4 | 13 | 12 | 110 |
| LD | 11 | 12 | 14 | 17 | 21 | 23 | 18 | 22 | 18 | 22 | 17 | 193 |
| Maternal | 0 | 0 | 0 | 0 | 0 | 1 | 0 | 15 | 0 | 36 | 0 | 52 |
| Medical harms | 576 | 172 | 66 | 15 | 7 | 6 | 8 | 7 | 10 | 8 | 14 | 890 |
| Meningitis | 1,413 | 1,050 | 302 | 80 | 21 | 19 | 20 | 35 | 28 | 22 | 23 | 3,014 |
| Neonatal / prematurity | 690,238 | 55,936 | 2,927 | 544 | 384 | 370 | 398 | 361 | 390 | 354 | 381 | 752,284 |
| Neuro devel | 110 | 110 | 110 | 111 | 131 | 63 | 223 | 98 | 276 | 161 | 376 | 1,770 |
| Neurological | 242 | 180 | 440 | 75 | 40 | 59 | 66 | 79 | 148 | 161 | 219 | 1,707 |
| Nutrition | 170 | 170 | 175 | 108 | 313 | 90 | 189 | 238 | 43 | 272 | 43 | 1,811 |
| Oral | 0 | 0 | 0 | 4 | 54 | 67 | 58 | 87 | 74 | 124 | 103 | 571 |
| Other cancer | 1,728 | 298 | 92 | 104 | 74 | 75 | 72 | 122 | 131 | 169 | 180 | 3,047 |
| Other unintentional injuries | 609 | 248 | 170 | 196 | 242 | 289 | 379 | 355 | 544 | 421 | 717 | 4,169 |
| Pain | 0 | 0 | 0 | 0 | 63 | 346 | 296 | 819 | 616 | 1,513 | 1,116 | 4,768 |
| Pneumonia | 8,049 | 2,312 | 734 | 107 | 25 | 22 | 20 | 26 | 29 | 35 | 40 | 11,399 |
| Rare ID | 1,776 | 1,780 | 163 | 33 | 32 | 19 | 24 | 35 | 23 | 44 | 28 | 3,956 |
| Road injuries | 167 | 93 | 45 | 66 | 70 | 101 | 140 | 248 | 613 | 252 | 822 | 2,617 |
| Self harm | 0 | 0 | 0 | 0 | 0 | 10 | 17 | 145 | 402 | 249 | 903 | 1,726 |
| SIDS |  | 4,109 | 2,002 |  |  |  |  |  |  |  |  | 6,111 |
| Skin | 3 | 16 | 100 | 999 | 879 | 776 | 595 | 689 | 482 | 652 | 448 | 5,637 |
| STD | 3,661 | 419 | 47 | 1 | 0 | 2 | 1 | 12 | 12 | 22 | 21 | 4,198 |
| Subs use | 0 | 0 | 0 | 0 | 0 | 16 | 25 | 552 | 1,035 | 878 | 1,962 | 4,469 |
| Violence | 525 | 117 | 58 | 27 | 19 | 35 | 31 | 67 | 70 | 79 | 112 | 1,141 |
| Vision | 12 | 13 | 23 | 59 | 85 | 87 | 87 | 83 | 83 | 80 | 81 | 691 |
| Wilms | 50 | 12 | 14 | 15 | 9 | 6 | 4 | 4 | 3 | 5 | 4 | 127 |
| **Grand Total** | **946,214** | **100,711** | **13,023** | **4,444** | **4,416** | **6,219** | **5,933** | **10,305** | **9,689** | **12,921** | **12,456** | **1,126,331** |

### Table A4. DALYS per 100,000 in 2007 and 2017 and change in decade 1998-2007 and 2008-2017 by age and sex

|  |  | Early Neonatal | | Late Neonatal | | Infant | | 1 to 4 | | 5 to 9 | | 10 to 14 | | 15 to 19 | | 20 to 24 | |
| --- | --- | --- | --- | --- | --- | --- | --- | --- | --- | --- | --- | --- | --- | --- | --- | --- | --- |
| Cause Name |  | 2007 | 2017 | 2007 | 2017 | 2007 | 2017 | 2007 | 2017 | 2007 | 2017 | 2007 | 2017 | 2007 | 2017 | 2007 | 2017 |
| All causes | % change in decade | -14 | -18 | -11 | -17 | -15 | -21 | -9 | -8 | -9 | -5 | -4 | -3 | -5 | -7 | -6 | -6 |
|  | DALYS per 100,000 | 1152,536 | 943673 | 120322 | 100433 | 16,464 | 12,984 | 4,836 | 4,442 | 4,630 | 4,412 | 6,279 | 6,076 | 10,717 | 9,997 | 13,518 | 12,689 |
| Neonatal | % change in decade | -18 | -18 | -15 | -18 | 10 | -15 | -2 | 8 | 1 | 13 | 2 | 11 | 2 | 10 | 3 | 10 |
|  | DALYS per 100,000 | 844,193 | 688261 | 67,460 | 55,724 | 3,407 | 2,915 | 504 | 544 | 340 | 384 | 345 | 384 | 340 | 376 | 333 | 368 |
| Congenital | % change in decade | 1 | -17 | 5 | -15 | -4 | -22 | -8 | -14 | -4 | -7 | -4 | -7 | -5 | -7 | -3 | -5 |
|  | DALYS per 100,000 | 271,269 | 224499 | 33,796 | 28,661 | 4,280 | 3,324 | 557 | 478 | 268 | 251 | 257 | 240 | 285 | 264 | 262 | 248 |
| SIDS | % change in decade |  |  | -37 | -17 | -43 | -23 |  |  |  |  |  |  |  |  |  |  |
|  | DALYS per 100,000 |  |  | 4,970 | 4,103 | 2,599 | 1,995 |  |  |  |  |  |  |  |  |  |  |
| LRTI | % change in decade | -16 |  | -28 | -18 | -36 | -29 | -25 | -22 |  |  |  |  |  |  |  |  |
|  | DALYS per 100,000 | 10,282 |  | 2,787 | 2,304 | 1,035 | 731 | 137 | 107 |  |  |  |  |  |  |  |  |
| Other unspecified infectious diseases | % change in decade |  |  | 38 | -13 | 2 | -20 |  |  |  |  |  |  |  |  |  |  |
|  | DALYS per 100,000 |  |  | 2,005 | 1,744 | 177 | 141 |  |  |  |  |  |  |  |  |  |  |
| Drug use disorders | % change in decade |  |  |  |  |  |  |  |  |  |  |  |  | 2 | 3 | 7 | -4 |
|  | DALYS per 100,000 |  |  |  |  |  |  |  |  |  |  |  |  | 645 | 650 | 1,104 | 1,007 |
| Headache | % change in decade |  |  |  |  |  |  |  |  | 0 | 0 | 0 | -1 | 0 | 0 | 0 | 0 |
|  | DALYS per 100,000 |  |  |  |  |  |  |  |  | 92 | 92 | 494 | 491 | 877 | 874 | 1,204 | 1,203 |
| Endocrine | % change in decade |  |  | 39 | -10 | 15 | -16 | 14 | -11 | -1 | -16 | 0 | -14 |  |  |  |  |
|  | DALYS per 100,000 |  |  | 2,269 | 2,047 | 447 | 373 | 134 | 119 | 74 | 62 | 71 | 61 |  |  |  |  |
| back pain | % change in decade |  |  |  |  |  |  |  |  | -1 | 1 | 0 | 1 | -2 | 1 | -7 | -2 |
|  | DALYS per 100,000 |  |  |  |  |  |  |  |  | 55 | 55 | 280 | 284 | 611 | 617 | 1,084 | 1,061 |
| Depression | % change in decade |  |  |  |  |  |  |  |  |  | 5 | -4 | 6 | -6 | 3 | -6 | -1 |
|  | DALYS per 100,000 |  |  |  |  |  |  |  |  |  | 42 | 277 | 294 | 608 | 626 | 789 | 782 |
| Gynecological | % change in decade |  |  |  |  |  |  |  |  |  |  |  |  | 3 | -1 | 2 | 1 |
|  | DALYS per 100,000 |  |  |  |  |  |  |  |  |  |  |  |  | 314 | 311 | 655 | 659 |
| Dermatitis | % change in decade |  |  |  |  |  |  | 2 | -2 | 1 | -5 | -1 | -8 | 0 | -7 | 1 | -3 |
|  | DALYS per 100,000 |  |  |  |  |  |  | 730 | 716 | 600 | 569 | 456 | 421 | 345 | 323 | 264 | 256 |
| Self-harm | % change in decade |  |  |  |  |  |  |  |  |  |  |  |  | -20 | -11 | -26 | -7 |
|  | DALYS per 100,000 |  |  |  |  |  |  |  |  |  |  |  |  | 317 | 274 | 657 | 576 |
| Anxiety | % change in decade |  |  |  |  |  |  |  |  | -2 | 1 | -2 | 1 | -1 | 1 | 0 | 0 |
|  | DALYS per 100,000 |  |  |  |  |  |  |  |  | 222 | 224 | 509 | 515 | 563 | 568 | 502 | 502 |
| Meningitis | % change in decade |  |  | -41 |  | -43 | -44 | -44 | -39 |  |  |  |  |  |  |  |  |
|  | DALYS per 100,000 |  |  | 1,247 |  | 447 | 252 | 107 | 65 |  |  |  |  |  |  |  |  |
| Asthma | % change in decade |  |  |  |  |  |  | -8 | -2 | -11 | 0 | -6 | -9 | -1 | -15 | 1 | -19 |
|  | DALYS per 100,000 |  |  |  |  |  |  | 279 | 273 | 360 | 356 | 407 | 370 | 482 | 408 | 486 | 394 |
| Conduct | % change in decade |  |  |  |  |  |  |  |  | -1 | -9 | 2 | 2 | 1 | 1 |  |  |
|  | DALYS per 100,000 |  |  |  |  |  |  |  |  | 202 | 184 | 458 | 465 | 347 | 350 |  |  |
| Road injuries | % change in decade |  |  |  |  |  |  | -35 | -39 | -43 | -35 | -38 | -35 | -29 | -42 | -25 | -32 |
|  | DALYS per 100,000 |  |  |  |  |  |  | 100 | 60 | 100 | 63 | 167 | 108 | 740 | 406 | 784 | 502 |
| Alcohol use disorders | % change in decade |  |  |  |  |  |  |  |  |  |  |  |  | 3 | -2 | 6 | -1 |
|  | DALYS per 100,000 |  |  |  |  |  |  |  |  |  |  |  |  | 148 | 143 | 423 | 413 |
| Bipolar | % change in decade |  |  |  |  |  |  |  |  |  |  | -1 | 0 | -2 | 0 | -2 | 0 |
|  | DALYS per 100,000 |  |  |  |  |  |  |  |  |  |  | 102 | 102 | 339 | 339 | 343 | 343 |
| Acne | % change in decade |  |  |  |  |  |  |  |  |  |  | 17 | 9 | 17 | 10 | 15 | 11 |
|  | DALYS per 100,000 |  |  |  |  |  |  |  |  |  |  | 126 | 137 | 406 | 448 | 200 | 222 |
| Motor neuron disease | % change in decade |  |  |  |  | -10 |  |  |  |  |  |  |  |  |  |  |  |
|  | DALYS per 100,000 |  |  |  |  | 232 |  |  |  |  |  |  |  |  |  |  |  |
| Diarrhoea | % change in decade |  |  |  |  | 45 | -32 | -4 | -11 | 0 | 6 |  |  |  |  |  |  |
|  | DALYS per 100,000 |  |  |  |  | 534 | 366 | 150 | 134 | 52 | 55 |  |  |  |  |  |  |
| Falls | % change in decade |  |  |  |  |  |  | 4 | 0 | 5 | 4 | 3 | 2 | 2 | 0 | 1 | 0 |
|  | DALYS per 100,000 |  |  |  |  |  |  | 92 | 92 | 134 | 140 | 199 | 204 | 269 | 269 | 333 | 332 |
| Foreign body | % change in decade |  |  |  |  | -5 | -24 | -11 | -19 |  |  |  |  |  |  |  |  |
|  | DALYS per 100,000 |  |  |  |  | 368 | 282 | 61 | 50 |  |  |  |  |  |  |  |  |
| Other neurological disorders | % change in decade |  |  |  |  | -6 | 14 | -25 | 11 |  |  |  |  |  |  |  |  |
|  | DALYS per 100,000 |  |  |  |  | 293 | 335 | 57 | 63 |  |  |  |  |  |  |  |  |
| Eating disorders | % change in decade |  |  |  |  |  |  |  |  |  |  |  | 4 | 8 | 4 | 7 | 2 |
|  | DALYS per 100,000 |  |  |  |  |  |  |  |  |  |  |  | 58 | 185 | 192 | 244 | 248 |
| Neck pain | % change in decade |  |  |  |  |  |  |  |  |  |  |  |  | -14 | 1 | -19 | 3 |
|  | DALYS per 100,000 |  |  |  |  |  |  |  |  |  |  |  |  | 99 | 101 | 246 | 253 |
| URTI | % change in decade |  |  |  |  | -2 | -2 | -1 | -1 | 0 | -1 | 1 | -1 | 1 | -1 |  |  |
|  | DALYS per 100,000 |  |  |  |  | 189 | 186 | 191 | 189 | 170 | 169 | 130 | 130 | 115 | 114 |  |  |
| Other mental disorders | % change in decade |  |  |  |  |  |  |  |  |  |  |  |  |  |  | 0 | 0 |
|  | DALYS per 100,000 |  |  |  |  |  |  |  |  |  |  |  |  |  |  | 156 | 156 |
| Other musculoskeletal disorders | % change in decade |  |  |  |  |  |  |  |  |  |  |  |  |  |  | 0 | -7 |
|  | DALYS per 100,000 |  |  |  |  |  |  |  |  |  |  |  |  |  |  | 161 | 150 |
| Diabetes | % change in decade |  |  |  |  |  |  |  |  |  |  |  |  | 98 | -8 | 65 | 9 |
|  | DALYS per 100,000 |  |  |  |  |  |  |  |  |  |  |  |  | 109 | 101 | 197 | 213 |
| iron deficiency | % change in decade |  |  |  |  |  | 2 | -47 | 17 | -26 | 3 | 32 | -3 | -17 | -3 | -3 | -12 |
|  | DALYS per 100,000 |  |  |  |  |  | 122 | 66 | 76 | 264 | 255 | 129 | 125 | 176 | 129 | 154 | 145 |
| Viral skin diseases | % change in decade |  |  |  |  |  |  | 0 | 0 | 0 | 0 | 0 | 0 |  |  |  |  |
|  | DALYS per 100,000 |  |  |  |  |  |  | 134 | 134 | 177 | 177 | 101 | 101 |  |  |  |  |
| Epilepsy | % change in decade |  |  |  |  |  |  | -6 | 4 | -7 | 5 | -3 | -2 | 2 | -14 | 1 | -15 |
|  | DALYS per 100,000 |  |  |  |  |  |  | 91 | 95 | 91 | 95 | 111 | 108 | 166 | 143 | 204 | 172 |
| Cardiomyopathy and myocarditis | % change in decade |  |  |  |  | -25 | -34 | -27 |  |  |  |  |  |  |  |  |  |
|  | DALYS per 100,000 |  |  |  |  | 200 | 132 | 44 |  |  |  |  |  |  |  |  |  |
| Violence | % change in decade |  |  |  |  |  |  |  |  |  |  |  |  |  |  | -8 |  |
|  | DALYS per 100,000 |  |  |  |  |  |  |  |  |  |  |  |  |  |  | 124 |  |
| Age-related and other hearing loss | % change in decade |  |  |  |  |  |  |  |  |  |  |  |  |  |  |  | 10 |
|  | DALYS per 100,000 |  |  |  |  |  |  |  |  |  |  |  |  |  |  |  | 115 |
| ASD | % change in decade |  |  |  |  |  |  | 0 | 0 | 0 | 0 | 0 | 0 | 0 | 0 |  |  |
|  | DALYS per 100,000 |  |  |  |  |  |  | 107 | 107 | 105 | 105 | 102 | 102 | 99 | 99 |  |  |
| Blindness | % change in decade |  |  |  |  |  |  | -1 | -1 | -2 | -1 | -2 | -1 |  |  |  |  |
|  | DALYS per 100,000 |  |  |  |  |  |  | 59 | 59 | 85 | 85 | 88 | 87 |  |  |  |  |
| CNS cancer | % change in decade |  |  |  |  |  |  | 4 | -10 | -1 | -19 | -3 | -13 |  |  |  |  |
|  | DALYS per 100,000 |  |  |  |  |  |  | 80 | 72 | 89 | 73 | 67 | 58 |  |  |  |  |
| Other malignancies | % change in decade |  |  |  |  |  |  | -7 | -2 | -5 | -7 | 0 | -5 |  |  |  |  |
|  | DALYS per 100,000 |  |  |  |  |  |  | 90 | 88 | 67 | 63 | 64 | 61 |  |  |  |  |
| Leukemia | % change in decade |  |  |  |  |  |  | -14 | -21 | -31 | -28 | -31 |  |  |  |  |  |
|  | DALYS per 100,000 |  |  |  |  |  |  | 89 | 70 | 68 | 49 | 66 |  |  |  |  |  |
| Urticaria | % change in decade |  |  |  |  |  |  | 0 | 0 | 0 | 0 |  |  |  |  |  |  |
|  | DALYS per 100,000 |  |  |  |  |  |  | 76 | 76 | 45 | 45 |  |  |  |  |  |  |
| Oral disorders | % change in decade |  |  |  |  |  |  |  |  | -7 | -2 | -2 | -1 |  |  |  |  |
|  | DALYS per 100,000 |  |  |  |  |  |  |  |  | 55 | 54 | 63 | 63 |  |  |  |  |
| Fire, heat, and hot substances | % change in decade |  |  |  |  |  |  | -41 |  |  |  |  |  |  |  |  |  |
|  | DALYS per 100,000 |  |  |  |  |  |  | 58 |  |  |  |  |  |  |  |  |  |
| Vitamin A deficiency | % change in decade |  |  |  |  |  |  |  |  | -31 | -12 |  |  |  |  |  |  |
|  | DALYS per 100,000 |  |  |  |  |  |  |  |  | 60 | 52 |  |  |  |  |  |  |
| Drowning | % change in decade |  |  |  |  |  |  | -23 |  |  |  |  |  |  |  |  |  |
|  | DALYS per 100,000 |  |  |  |  |  |  | 54 |  |  |  |  |  |  |  |  |  |
| Otitis media | % change in decade |  |  |  |  |  |  | -3 | 0 |  | -1 |  |  |  |  |  |  |
|  | DALYS per 100,000 |  |  |  |  |  |  | 54 | 54 |  | 40 |  |  |  |  |  |  |

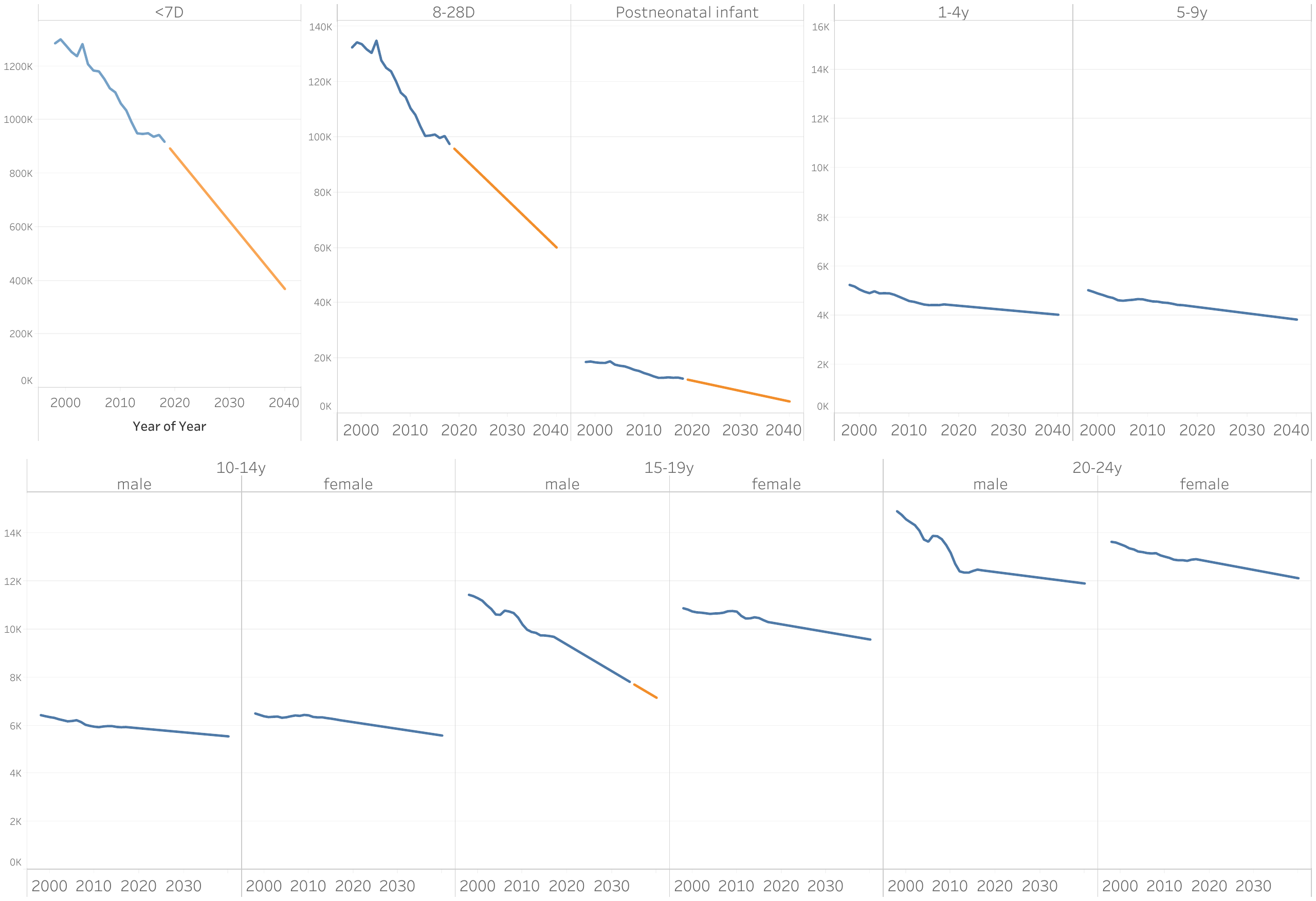

### Figure A2. Observed total DALYS per 100,000 from 1998 to 2017 and forecast from 2018 to 2040, by age

Note: Where future forecasts are significantly lower (or higher) than in 2017 (i.e. values fall outside the uncertainty intervals for 2017), these are highlighted in orange.
